# Clinical and Social Determinants of Guideline-Directed Medical Therapy Optimization in Heart Failure with Reduced Ejection Fraction

**DOI:** 10.64898/2026.11.02.26367137

**Authors:** Maxine Nelson, Marcus Kwon, Kevin Driscoll, Oroshay Kaiwan, Mark E. Dunlap, Khalill Murad, Anna Rybińska-Campbell, Douglas D. Gungzler, Phoo Pwint Nandar

## Abstract

**Background:** Heart failure with reduced ejection fraction (HFrEF) is associated with high morbidity and mortality. Guideline-directed medical therapy (GDMT) for HFrEF has been shown to improve outcomes, with higher GDMT scores at hospital discharge being associated with lower mortality, fewer rehospitalizations and reduced healthcare costs. Yet, disparities in the utilization of GDMT remain. In this study, we examined the effect of various clinical, demographic, and socioeconomic factors on the utilization of GDMT in HFrEF patients referred to Cardiology in a tertiary care safety net hospital, using a standardized GDMT scoring system.

**Methods:** We conducted a single-center cross-sectional retrospective analysis of patients who had an index HFrEF ≤ 40% diagnosis between January 1, 2024, to December 31, 2024, and who were referred to Cardiology at a tertiary care safety net hospital. Baseline characteristics, laboratory investigations, health care utilization, and GDMT score data were extracted at the ambulatory care visit nearest to 90 days from the index HF diagnosis. GDMT score ranges from 0 to 9 factored in doses of beta-blocker (BB), angiotensin-converting enzyme inhibitor/angiotensin receptor blocker (ACEi/ARB), mineralocorticoid receptor antagonist (MRA), sodium–glucose cotransporter 2 inhibitor (SGLT2i), angiotensin receptor–neprilysin inhibitor (ARNI), isosorbide dinitrate/hydralazine (ISDN/HYD). Multivariable logistic regression was performed to examine associations between low GDMT scores (0-4) nearest to 90 days from new onset HFrEF diagnosis and several clinical, demographic, and socioeconomic variables.

**Results:** In 200 patients meeting entry criteria, mean age was 62.7 years, 64.0% were male, 47.5% were classified as white and 42.0% as Black. 39.5% had a low GDTM score (<5). The following were less likely to be associated with low GDMT score: diabetes mellitus (adjusted OR: 0.41, 95%CI=0.18-0.91) and frequent medical appointments (adjusted OR: 0.37, 95% CI=0.25-0.54), while higher serum eGFR was more likely to be associated with low GDMT score (adjusted OR: 1.02, 95%CI=1.00-1.03). There was no significant association between GDMT score and age, sex, race, or payer status.

**Conclusion:** In patients with HFrEF referred to Cardiology in our safety net tertiary care center, higher utilization of GDMT at 90 days was achieved in those who have diabetes mellitus, more frequent medical appointments, or lower serum eGFR. There was no difference in GDMT utilization according to age, sex, race, or payer status predictive of lower GDMT scores. Contrary to findings from previous studies, no significant differences were observed across demographic or payer groups, emphasizing the role of healthcare access and utilization.

## Introduction

Multiple analyses of diverse heart failure (HF) populations have shown that patients on optimized heart failure regimens experience significantly lower rates of death and HF-related hospitalizations.^1,2^ To capture the complexity of HF treatment, the guideline-directed medical therapy (GDMT) score created by the Heart Failure Collaboratory and the Academic Research Consortium was implemented.^3^ Martyn, T. et al. also described a scoring system based on previously proposed GDMT scores.^4^

Higher hospital GDMT scores at discharge correlates with better prognosis in HFrEF and mildly reduced ejection fraction (HFmrEF) with reduced mortality, lower rehospitalization rates and reduced costs of healthcare, underscoring the importance of optimizing GDMT to improve clinical outcomes and reduce costs in HFrEF management.^3^

Aiming for a simple GDMT score of at least 5 may still improve patient outcomes, if all four cornerstone medications cannot be prescribed.^3^ However, disparities in GDMT implementation persist, and this study aims to identify these gaps, i.e. race, payer status, and numbers of office visits.

## Methods

We conducted a single-center cross-sectional retrospective analysis of cardiology patients with an index HFrEF ≤40% diagnosis between January 1, 2024, to December 31, 2024 at a tertiary care safety net hospital. The study was approved by the Institutional Review Board, and the requirement for informed consent was waived. Exclusion criteria included eGFR <20 mL/min/1.73 m^2^ or on dialysis, prior or planned advanced heart failure therapies (left ventricular assist device [LVAD] or Orthotopic Heart Transplantation [OHT]), stage D heart failure under evaluation for advanced therapies, hospice enrollment, secondary cardiomyopathies (including infiltrative, tachycardia-mediated, post transcatheter aortic valve replacement (TAVR), or muscular dystrophy–related), or severe valvular disease.

Comprehensive data were extracted from the electronic medical record by manual chart review using a standardized data collection form by four members of the research team who were trained in the study protocol. Data including baseline characteristics (age, sex, race), laboratory investigations (eGFR), health care utilization (payor status, number of visits) and GDMT score data were extracted at the ambulatory care visit nearest to 90 days from the index HF diagnosis. Complete data was available for all 200 patients.

The GDMT score (range 0-9) factored doses of RAS inhibitors were scored 0 if not initiated, 1 if <50% of target dose, and 2 if 50–100% of target dose; ARNIs were scored 2 regardless of dose; BBs were scored 0 if not initiated, 1 if <50% of target dose, and 2 if 50–100% of target dose; and MRAs and SGLT2 inhibitors were scored 0 if not initiated and 2 if initiated regardless of dose and ISDN/HYD, scored 1for any dose if used in combination.

Data were tabulated and multivariable logistic regression was performed to examine associations between low GDMT score (0-4) at 90 days from index HFrEF diagnosis and demographic, clinical and healthcare utilization variables. Statistical analyses were conducted using Stata 18 (Stata Corp. LLC, College Station, TX, USA).

## Results

The mean age was 62.7 ± 12.7 years, and the mean estimated glomerular filtration rate (eGFR) was 71.7 ± 25.0 mL/min/1.73 m^2^. The cohort was predominantly male (64.0%, n = 128). Nearly half of the participants were white (47.5%, n = 95), followed by Black (42.0%, n = 84), while 10.5% (n = 12) were another race. 63% of patients had private insurance, 26.5% had public insurance, and 10.5% were uninsured. Regarding outpatient follow-up, the majority of patients attended two (35.5%, n = 71) or three (29.5%, n = 59) office visits during the study period. Fewer patients had one visit (15.5%, n = 31), four visits (18.0%, n = 36), or five visits (1.5%, n = 3). (*Table 1*)

**Table 1.** Baseline characteristics of the cohort.

| **Characteristics** |  |
|---|---|
| N | 200 |
| Age, years *Mean (SD*) | 62.7 ± 12.7 |
| Serum eGFR, mL/min/1.73 m² *Mean (SD)* | 71.7 ± 25.0 |
| Sex, *n* (%) |  |
| Male sex *n* (%) | 128 (64.0%) |
| Female sex *n* (%) | 72 (36.0%) |
| Race, *n* (%) |  |
| White race *n* (%) | 95 (47.5%) |
| Black race *n* (%) | 84 (42.0%) |
| Other race *n* (%) | 21 (10.5%) |
| Payor type, *n* (%) |  |
| Private insurance | 126 (63.0%) |
| Public insurance | 53 (26.5%) |
| Uninsured | 21 (10.5%) |
| GDMT score total, *n* (%) |  |
| GDMT SCORE 0-4 | 79 (39.5%) |
| GDMT score 5-9 | 121 (60.5%) |

eGFR, estimated glomerular filtration rate; GDMT, guideline-directed medical therapy; HF, heart failure.

In the multivariable logistic regression model adjusting for demographic and clinical covariates, diabetes mellitus, serum eGFR, and the number of outpatient office visits were associated with a low GDMT score (*Table 2, Figure 1*). Patients with diabetes had 59.0% lower odds of a low GDMT score compared with those without diabetes (adjusted OR 0.41, 95% CI 0.18–0.91, *p* < 0.05). Higher serum eGFR remained associated with increased odds of a low GDMT score (adjusted OR 1.02, 95% CI 1.00–1.03, *p* < 0.05). Outpatient office visits following discharge were significantly associated with GDMT scare, with each additional outpatient office visit associated with a 63.0% reduction in the odds of a low GDMT score (adjusted OR 0.37, 95% CI 0.25 – 0.54, *p* < 0.001).

**Table 2.** Multivariable logistic regression of variables associated with a guideline-directed medical therapy score of 4 or lower.

| **Variable** | **Adjusted Odds Ratio (95% CI)** | **P value** |
|---|---|---|
| Age (per year) | 1.01 (0.98–1.05) | 0.338 |
| Male vs Female | 0.88 (0.44–1.77) | 0.723 |
| Black/African American vs White | 1.11 (0.54–2.30) | 0.774 |
| Other Race vs White | 1.66 (0.53–5.20) | 0.385 |
| Atrial Fibrillation within 1 Year Following the GDMT Score Assessment | 1.05 (0.48–2.27) | 0.904 |
| Diabetes Mellitus | 0.41 (0.18–0.91) | 0.029 |
| Serum eGFR (per mL/min/1.73 m²) | 1.02 (1.00–1.03) | 0.021 |
| HF hospitalization within 1 Year Following the GDMT Score Assessment | 0.56 (0.28–1.12) | 0.103 |
| Number of Office Visits | 0.37 (0.25–0.54) | <0.001 |

eGFR, estimated glomerular filtration rate; GDMT, guideline-directed medical therapy; HF, heart failure;

**Figure 1.**
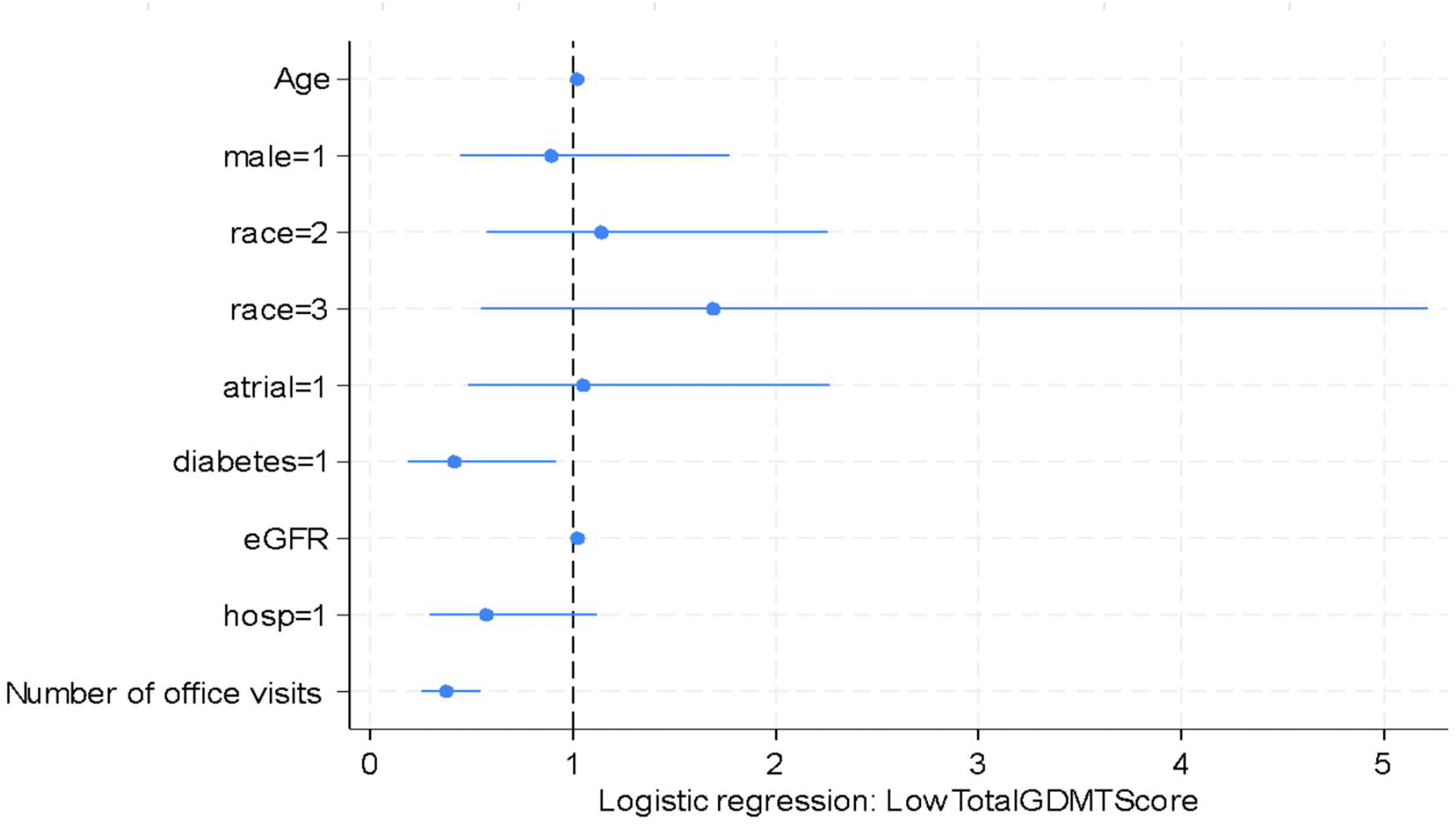
Forest Plot of Predictors of GDMT Score less than 4.

## Discussion

In this single-center retrospective cohort study of patients with HFrEF, we evaluated clinical and social patient-level factors associated with the achievement of GDMT using a standardized GDMT score nearest to 90 days following index HF diagnosis. Contrary to our initial hypothesis, demographic characteristics including age, sex, and race were not independently associated with lower GDMT utilization after multivariable adjustment. Instead, follow up office visits, renal function, and diabetes mellitus were the primary predictors of GDMT implementation regardless of demographic characteristics.

There was a strong association between outpatient follow up and GDMT optimization. Patients with an increased number of office visits had considerably lower odds of a low GDMT score. This observation reinforces the concept that optimization of GDMT is a dynamic process that requires regular patient reassessment, medication titration, laboratory monitoring, and patient education instead of reliance on a single clinical encounter. Consistent with previous studies, we found that dedicated heart failure clinics and structured follow up programs improved earlier achievement of target medication doses.^5^ Our findings further emphasize the importance of frequent outpatient follow up as a modifiable strategy to optimize GDMT in patients with HFrEF.

Patients with diabetes mellitus were less likely to have a low GDMT score. Generally, patients with diabetes have more frequent follow-ups with healthcare providers, thereby providing an increased opportunity for GDMT optimization. Furthermore, the implementation and widespread adaptation of SGLT2i for HFrEF and diabetes has contributed to optimized GDMT in this population.

We observed that higher serum eGFR was associated with slightly higher odds of a low GDMT score. Though statistically significant, the magnitude of the association was modest, suggesting limited clinical impact. This finding is unexpected and may be a reflection of unmeasured differences in clinical practice or residual confounding. Patients with chronic kidney disease (CKD) have elevated cardiovascular risk and would often undergo more frequent cardiovascular follow-ups, providing increased opportunities for initiation and titration of medications.

Furthermore, other clinical factors influence GDMT prescription beyond renal function, including electrolyte abnormalities, blood pressure, medication tolerance, and clinical judgment, which were not captured in this analysis.

Unlike what has been previously reported in national registries, lower GDMT utilization among women, racial groups and socioeconomically disadvantaged patients was not observed in this cohort. This may reflect effectively implementing standardized heart failure management protocols at our institution. Regardless, the absence of these observed disparities is reassuring and is suggestive that standardized care pathways may help with the mitigation of inequities observed in HFrEF management.

Our findings indicate that health care utilization may be a more significant determinant of GDMT optimization than demographic characteristics. Frequent outpatient follow-up represents an important quality improvement target for the opportunity to improve GDMT. The expansion of multidisciplinary heart failure care through pharmacist-led medication titration remote monitoring and telemedicine may provide increased opportunities for the initiation, titration of evidence-based therapies while providing patient education for HFrEF. A standardized electronic health record (EHR) dot phrase could further facilitate real-time identification of GDMT gaps and prompt clinicians to initiate or intensify therapy when appropriate. Furthermore, implementation of a pre-discharge GDMT score could facilitate identification of patients requiring additional intervention. Specifically, a GDMT score <5 at the time of discharge could trigger an inpatient clinical pharmacist consultation or referral to a dedicated outpatient heart failure clinic for medication optimization and longitudinal follow-up. This standardized approach may help identify therapeutic gaps before discharge and provide a structured pathway for continued GDMT optimization after hospitalization.

This study is limited by its single-center design and retrospective nature but reflects clinical practices of a large volume tertiary care safety net hospital. The GDMT score was assessed at an approximate 90-day HFrEF diagnosis and may not reflect subsequent medication optimization or long-term adherence. Furthermore, since GDMT assessment was performed at an approximate 90-day interval, patients may have had variable durations available for medication initiation and titration, which could have influenced their final GDMT scores.

## Data Availability

The data underlying this study are not publicly available due to patient confidentiality and institutional privacy restrictions.

